# Epidemiology of *Shigella sonnei* and *Shigella flexneri* in the Global Pediatric Diarrhea Surveillance network, 2017–2022

**DOI:** 10.64898/2026.09.14.26363009

**Authors:** Jie Liu, Myra Lewontin, Sarah E. Elwood, Shilpa S. Iyer, Sébastien Antoni, Gloria Rey-Benito, Claudia Ortiz, Goitom Weldegebriel, Joseph Biey, Jason M. Mwenda, Roberta Pastore, Dovile Videbaek, Simarjit Singh, Emmanuel Njambe, Lucky Sangal, Deepak Dhongde, Varja Grabovac, Josephine Logronio, Kamal Fahmy, Amany Ghoniem, George Armah, Francis E. Dennis, Mapaseka L. Seheri, Nonkululeko Magagula, Kebareng Rakau-Nondela, Tulio M. Fumian, Irene T.A. Maciel, Elena Samoilovich, Galina Semeiko, Tintu Varghese, Sarah Thomas, Julie E. Bines, Dandi Li, Furqan Kabir, Eric R. Houpt, Mick N. Mulders, Heidi M. Soeters, James A. Platts-Mills, the Global Pediatric Diarrhea Surveillance network

## Abstract

**Background:** *Shigella* is a leading cause of diarrhea in children in low- and middle-income countries (LMICs). Optimizing the impact of vaccines necessitates an accurate characterization of *Shigella* serotype prevalence. We describe the burden, epidemiology and characteristics of *S. sonnei* and *S. flexneri* diarrhea in the Global Pediatric Diarrhea Surveillance (GPDS) network.

**Methods:** GPDS is a World Health Organization-coordinated surveillance network investigating the etiology of hospitalized diarrhea among children aged <5 years. GPDS enrolls children hospitalized with diarrhea at 38 sentinel surveillance sites in 31 LMICs. Randomly selected stool specimens from these children were tested by quantitative polymerase chain reaction to detect a broad range of enteropathogens as well as to identify *S. sonnei* and *S. flexneri* serotypes. We estimated pathogen-specific attributable fractions and incidence of diarrheal hospitalizations.

**Results:** During 2017–2022, GPDS enrolled 70,750 children aged <5 years hospitalized with diarrhea, of which 16,458 (23.3%) were randomly selected for qPCR testing. Globally, *Shigella* caused 9.8% (95% Confidence Interval [CI]: 8.4, 11.3) of hospitalized diarrhea, with 28.3% of cases occurring in children aged ≤12 months. The estimated attributable incidence of *Shigella*-associated diarrhea hospitalizations decreased from 1.27 (95% CI: 0.95, 1.64) per 1,000 child years in 2017–2018 to 0.88 (0.67, 1.10) in 2021–2022. Among typeable *Shigella* cases, 34.8% were *S. sonnei*, 24.2% were *S. flexneri* 2a, and 9.9% were *S. flexneri* 1b. Vaccines currently in development would cover an estimated 59.0% to 72.2% of cases.

**Conclusions:** These findings support the role of *Shigella* as an important cause of the most severe diarrhea in young children. The breadth of homotypic protection provided by *Shigella* vaccines in clinical development varied by region, and these broadly representative data can aid in prioritization. Non-dysenteric *Shigella* could not be readily differentiated from other causes of hospitalized diarrhea based on clinical characteristics.

## Introduction

*Shigella* is a leading cause of acute diarrhea in children in low- and middle-income countries (LMICs) (1–6), causing an estimated 76,400 deaths among children aged <5 years globally in 2023 (5). In addition to causing severe disease and death among children, *Shigella* is also responsible for significant long term health consequences, including growth faltering and delayed cognitive development, as well as financial burdens to families and health systems (7). *Shigella* is comprised of four species – *Shigella flexneri*, *Shigella dysenteriae*, and *Shigella boydii*, each of which consists of multiple serotypes, and *Shigella sonnei*, which contains a single serotype (8). *S. flexneri* and *S. sonnei* are the dominant causes of endemic shigellosis in LMICs (4,9,10). While *S. flexneri* has been associated with a higher risk of dysentery than *S. sonnei*, the relative associations with other clinical characteristics including vomiting and dehydration are less clear (4,10).

Several *Shigella* vaccines are in development, the most advanced of which are subunit vaccines targeting the most common serotypes of *S. sonnei* and *S. flexneri* (11–14). Therefore, optimizing the potential impact of vaccination against *Shigella* necessitates an accurate characterization of global *Shigella* serotype prevalence. Most existing data on *Shigella* serotype distribution in LMICs comes from a limited set of research settings and from cultured *Shigella* isolates (1,4). However, *Shigella* culture is insensitive and not widely available, limiting the representativeness of the available serotyping data. Further, it is possible that *S. sonnei* culture recovery is higher, as this species is more adept at surviving nutrient starvation, maintaining extracellular viability, and outcompeting commensal flora (15,16). Consequently, estimates of *Shigella* species distributions based solely on culture may be biased. Techniques have emerged to use quantitative polymerase chain (qPCR) directly on stool samples to identify *S. sonnei* and *S. flexneri* serotypes (17,18,9), allowing for better characterization of the global distribution and prevalence of these dominant species.

We applied this qPCR serotyping approach to stool samples collected via the World Health Organization (WHO)-coordinated Global Pediatric Diarrhea Surveillance (GPDS) network (19). GPDS was established in 2017 as a public health surveillance network to investigate the etiology of hospitalized diarrhea among a broadly representative set of children aged <5 years in LMICs (3,19). The GPDS network enrolls children hospitalized with diarrhea at 38 sentinel surveillance sites in 31 countries, many with high *Shigella* burden, providing the basis for a more globally representative characterization of *Shigella* species and serotypes. In GPDS, stool samples are analyzed via a TaqMan Array card (TAC) qPCR assay for 16 enteric pathogens, including *Shigella*. The TAC qPCR cards also included targets to identify *Shigella* serotypes (17). Importantly, GPDS enrolls children aged <5 years in LMICs, which is the priority target population that WHO has identified for a *Shigella* vaccine (14,20). In this analysis, we describe the epidemiology and clinical characteristics of *S. sonnei* and *S. flexneri* in the GPDS network.

## Methods

Detailed methods for GPDS are described elsewhere (19). Briefly, GPDS sentinel surveillance hospitals enrolled a minimum of 100 cases per year of hospitalized diarrhea in children aged 0-59 months. GPDS sites prospectively enrolled all children admitted for diarrhea, regardless of symptom duration or the presence of blood in the stool. Diarrhea was defined as 3 or more loose stools in a 24-hour period. An acute diarrhea episode was defined as an illness with a duration prior to enrollment of fewer than 14 days (through a threshold of <7 days was used in some participating countries), while longer episodes were considered persistent.

Thirty-eight sentinel surveillance sites in 31 countries across all 6 WHO regions participated in GPDS in 2017 through 2022 (19). From each surveillance site, 100 cases were randomly selected for qPCR testing and were shipped to a Regional Reference Laboratory. Samples were tested using custom-designed TAC cards (Thermo Fisher, Waltham, MA, USA) (19), which included qPCR assays for the 16 enteric pathogens that were associated with pediatric diarrhea in two prior multisite studies that incorporated diarrheal cases and non-diarrheal controls: the Global Enteric Multicenter Study (GEMS) and the Etiology, Risk Factors, and Interactions of Enteric Infections and Malnutrition and the Consequences for Child Health and Development (MAL-ED) birth cohort study (1,2). Detections with a cycle threshold (Ct) value <35 were considered positive.

For analysis, we applied inverse probability of selection weights such that cases selected for testing were representative of all enrolled cases, including appropriately capturing seasonal variation in diarrhea etiology (19). As asymptomatic identification of enteropathogens is common in LMICs, especially when using molecular testing methods, we attributed diarrhea to specific enteropathogens based on pathogen quantity using models developed from qPCR re-analyses of the GEMS case-control study and the MAL-ED birth cohort study (1,2,21). Weighted population attributable fractions (AFs) for each pathogen were calculated and then applied to national-level estimates of both diarrhea hospitalizations and children aged <5 years from the Global Burden of Disease (GBD) 2021 study to estimate the pathogen-specific incidence of hospitalized diarrhea (22). Estimates were produced at the country, regional, and global levels in two-year analytic groupings (2017-2018, 2019-2020, and 2021-2022) and overall for the entire 2017–2022 period for select estimates.

To detect *Shigella*, the TAC qPCR cards included the *ipaH* gene target (19). Though *ipaH* can be found in both *Shigella* and enteroinvasive *Escherichia coli*, based on previously reported evidence that the vast majority of *ipaH* detections in these settings represent *Shigella*, we refer to detection of *ipaH* as *Shigella* throughout this manuscript (1,23). For cases where multiple pathogens were detected in the stool sample, we determined the single most likely etiologic pathogen by calculating the individual etiologic attributable fraction (AFe) for each pathogen within each diarrheal episode, serving as an episode- and pathogen quantity-specific metric of attribution of that pathogen as the cause of the illness. We then assigned attribution to the pathogen with the highest AFe (if the AFe was >0.2). Therefore, for this analysis, *Shigella*-attributable diarrhea was defined as the detection of *ipaH* with a *Shigella* AFe >0.2 and no other pathogen with a higher AFe. Starting in 2019, ten additional targets were added to the GPDS TAC qPCR cards to identify *S. flexneri* serotypes and *S. sonnei*; *S. flexneri* serotypes were assigned based on these targets using a refined algorithm, as previously described (18).

To calculate age-specific cumulative proportions and *Shigella* serotype distributions for geographic groupings and overall, we used a two-stage weighting approach that incorporated selection weights as well as the estimated number of attributable *Shigella* diarrhea hospitalizations in each country. First, we estimated country-specific distributions using patient selection weights, and then aggregated these distributions, weighted by the estimated number of attributable *Shigella* hospitalizations for each country, to calculate regional and overall cumulative proportions by age and serotype distributions. Homotypic *Shigella* vaccine coverage was estimated as the cumulative weighted proportion of serotypes included in proposed bivalent (*S. sonnei* + *S. flexneri* 2a), quadrivalent (*S. sonnei* + *S. flexneri* 2a, 3a, 6), and Generalized Modules for Membrane Antigens (GMMA; *S. sonnei* + *S. flexneri* 1b, 2a, 3a) formulations relative to the total weighted *Shigella* burden (11).

We evaluated the clinical presentation of *Shigella*-attributable diarrhea with clinical characteristics modeled as a function of the *Shigella* AFe for each episode. Differences in continuous severity metrics (e.g., severity score (19), maximum daily episodes) were assessed using linear regression, while the presence of specific binary characteristics (e.g., bloody diarrhea, severe dehydration, intravenous [IV] rehydration) was analyzed using Poisson regression with a log link to estimate relative risks. Models were adjusted for study site and year. We further compared clinical outcomes for *Shigella* AFes for the subset of episodes caused by *S. flexneri*, *S. sonnei*, and untyped *Shigella* (which could include *S. boydii*, *S. dysenteriae*, and enteroinvasive *E. coli* as well as potentially unidentified *S. sonnei* and *S. flexneri* serotypes) with the *Shigella* AFes of non-*Shigella* diarrhea.

The seasonality of hospitalized shigellosis was modeled as previously described (19). Modeled seasonal curves were overlaid with regional climate data (mean monthly rainfall and temperature) to qualitatively assess the association between meteorological drivers and *Shigella* species distribution.

For all analyses, point estimates and 95% confidence intervals were derived from the median and 2.5th and 97.5th quantiles respectively of the estimate distributions. All analyses were conducted in R version 4.0.2 and were run using GPDS surveillance data available as of 16 January 2026.

### Ethical considerations

Surveillance activities that are part of GPDS are exempt from research ethical review by WHO, since they are considered public health surveillance (24). Authors had no access to information that could identify individual participants during or after data collection.

## Results

GPDS enrolled 70,750 children aged <5 years with hospitalized diarrhea from 2017 to 2022, 16,458 (23.3%) of which were randomly selected for qPCR testing (Table 1). Of those tested, 2,399 (14.6%) had *Shigella* detected, and 1,673 (10.2%) had diarrhea estimated to be attributable to *Shigella*. *Shigella*-attributable cases were 56.1% male, similar to among all qPCR-tested cases (58.2%). *Shigella*-attributable cases had an older age distribution than all qPCR-tested cases. South Asia and East and Southern Africa contributed the largest numbers of *Shigella*-attributable cases, whereas none of the pediatric diarrhea cases from East Asia tested with qPCR were attributable to *Shigella*. Nearly all (98.2%) of *Shigella* cases presented with acute diarrhea, 63.6% reported vomiting, and around 70% experienced dehydration and required rehydration treatment. Bloody diarrhea was seen in 15.8% of *Shigella* cases with reported data on this clinical characteristic, compared to only 6.7% of all enrolled cases. In-hospital death was rare: 0.5% for all enrolled cases and 0.3% for *Shigella*-attributable cases.

**Table 1.** Characteristics of children with *Shigella*-attributable diarrhea in the Global Pediatric Diarrhea Surveillance network, 2017-2022.

|  | All enrolled cases<br>N = 70750 | Tested by qPCR<br>N = 16458 (23.3) | *Shigella*-attributable<br>cases<br>N = 1673 (10.2) |
| --- | --- | --- | --- |
| *Demographics* |  |  |  |
| Male | 41005 (58.0) | 9578 (58.3) | 936 (56.1) |
| <12 months of age | 31848 (45.0) | 7237 (45.7) | 359 (25.3) |
| 12-23 months of age | 22791 (32.2) | 5300 (32.1) | 565 (34.0) |
| 24-59 months of age | 16111 (22.8) | 3921 (22.2) | 749 (40.8) |
| *Geographic Region* |  |  |  |
| Central America | 7827 (11.1) | 1142 (11.1) | 179 (17.8) |
| Central and Western Europe | 10404 (14.7) | 1331 (14.9) | 146 (19.6) |
| East and Southern Africa | 14070 (19.9) | 3164 (19.7) | 410 (22.1) |
| East Asia | 3598 (5.1) | 1584 (5.1) | 0 (0) |
| Eastern Europe | 5281 (7.5) | 900 (7.5) | 30 (3.0) |
| South America | 4565 (6.5) | 1989 (6.5) | 274 (9.0) |
| South Asia | 8006 (11.3) | 2407 (11.4) | 325 (14.6) |
| Southeast Asia and Oceania | 10166 (14.4) | 1780 (14.3) | 95 (5.2) |
| West Africa | 6833 (9.7) | 2161 (9.6) | 214 (8.5) |
| *Clinical Characteristics* |  |  |  |
| Acute diarrhea | 69758 (98.6) | 16218 (98.7) | 1644 (98.2) |
| Bloody diarrhea | 4097 (6.7) | 1016 (6.5) | 255 (15.8) |
| Vomiting | 46830 (68.4) | 10958 (68.9) | 1042 (63.6) |
| Dehydration | 40927 (75.3) | 8145 (76.4) | 856 (70.1) |
| Rehydration therapy given | 39998 (74.7) | 7914 (76.1) | 834 (69.0) |
| In-hospital deaths | 319 (0.5) | 90 (0.4) | 7 (0.3) |

Characteristics are shown as N (%). Counts are unweighted; percentages are weighted and calculated from the subset of participants with non-missing data for each characteristic.

Overall, 65.8% of *Shigella*-attributable cases occurred in children aged ≤24 months, with 29.0% occurring in children aged ≤12 months (Table 2). However, the age distribution varied widely by geographic region, ranging from 10.7% aged ≤24 months in Eastern Europe to 89.0% aged ≤24 months in West Africa. West Africa also had the highest proportion of *Shigella* cases among infants, with 20.9% of *Shigella-*attributable cases occurring among children aged <6 months.

**Table 2.**
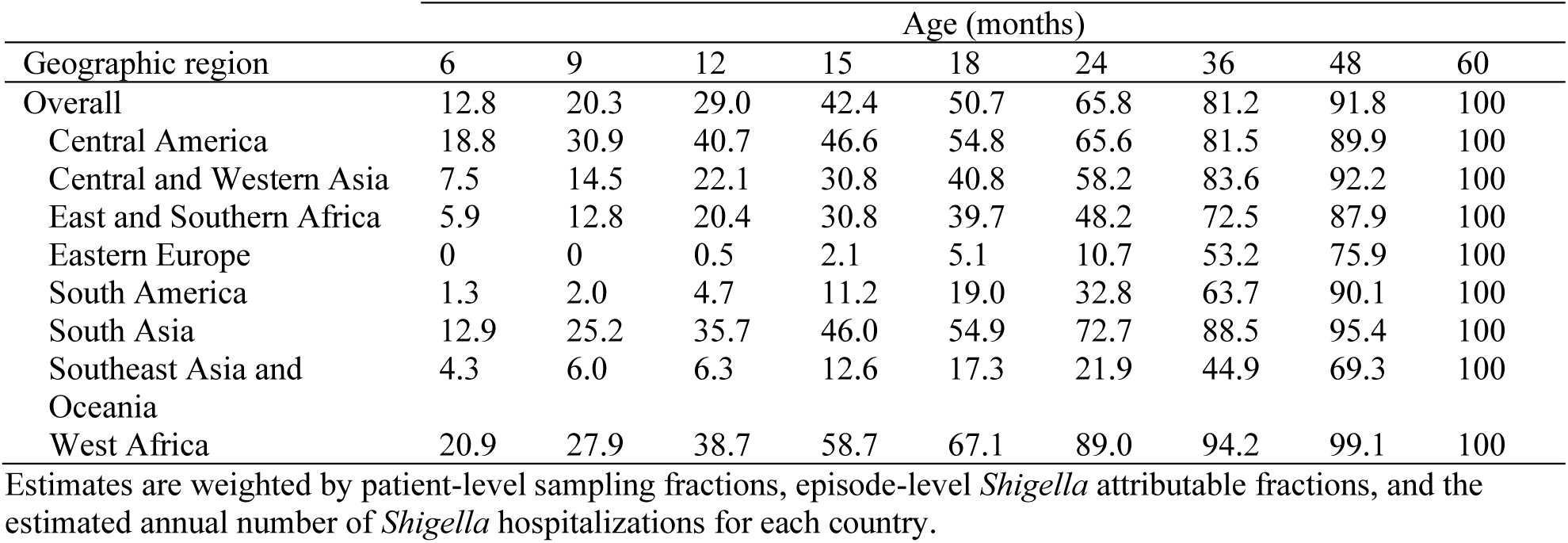
Weighted cumulative percent of children with *Shigella*-attributable diarrhea by age and geographic region in the Global Pediatric Diarrhea Surveillance network, 2017-2022.

| Geographic region | Age (months) |  |  |  |  |  |  |  |  |
| --- | --- | --- | --- | --- | --- | --- | --- | --- | --- |
|  | 6 | 9 | 12 | 15 | 18 | 24 | 36 | 48 | 60 |
| Overall | 12.8 | 20.3 | 29.0 | 42.4 | 50.7 | 65.8 | 81.2 | 91.8 | 100 |
| Central America | 18.8 | 30.9 | 40.7 | 46.6 | 54.8 | 65.6 | 81.5 | 89.9 | 100 |
| Central and Western Asia | 7.5 | 14.5 | 22.1 | 30.8 | 40.8 | 58.2 | 83.6 | 92.2 | 100 |
| East and Southern Africa | 5.9 | 12.8 | 20.4 | 30.8 | 39.7 | 48.2 | 72.5 | 87.9 | 100 |
| Eastern Europe | 0 | 0 | 0.5 | 2.1 | 5.1 | 10.7 | 53.2 | 75.9 | 100 |
| South America | 1.3 | 2.0 | 4.7 | 11.2 | 19.0 | 32.8 | 63.7 | 90.1 | 100 |
| South Asia | 12.9 | 25.2 | 35.7 | 46.0 | 54.9 | 72.7 | 88.5 | 95.4 | 100 |
| Southeast Asia and Oceania | 4.3 | 6.0 | 6.3 | 12.6 | 17.3 | 21.9 | 44.9 | 69.3 | 100 |
| West Africa | 20.9 | 27.9 | 38.7 | 58.7 | 67.1 | 89.0 | 94.2 | 99.1 | 100 |

Estimates are weighted by patient-level sampling fractions, episode-level *Shigella* attributable fractions, and the estimated annual number of *Shigella* hospitalizations for each country.

Overall, the proportion of all hospitalized diarrhea among children aged <5 years that was attributable to *Shigella* remained relatively constant at around 10% from 2017 to 2022 (Fig 1; S1 Table). In both East and Southern Africa and South Asia, the *Shigella* attributable fraction increased during this time period, while Central America and West Africa experienced decreases. The highest observed *Shigella* AF was in East and Southern Africa in 2021-2022 (18.4%; 95% Confidence Interval [CI]: 13.1, 24.8), whereas Eastern Europe had very low *Shigella* AFs (∼1%) and East Asia had no *Shigella* cases detected by GPDS during 2017-2022.

**Fig 1.**
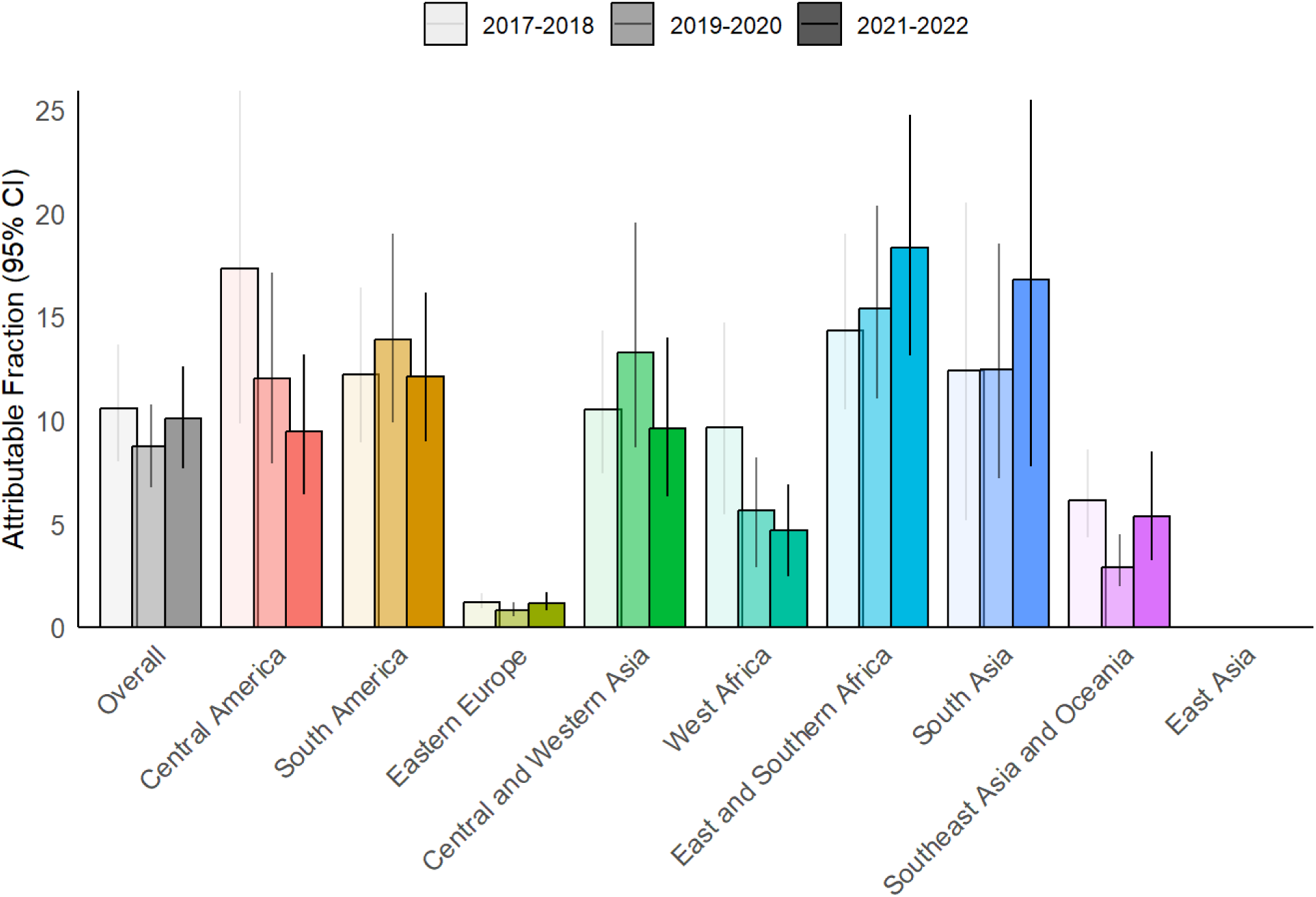
Attributable fractions of hospitalized diarrhea due to *Shigella* in children aged <5 years in 2017-2022 in the Global Pediatric Diarrhea Surveillance network both overall and by geographic region. Within each grouping, the attributable fractions were weighted by the site-level attributable incidence of hospitalized diarrhea. Bar shadings indicate the two-year periods of surveillance. Attributable fractions are expressed as a percent. No bars are included for East Asia, as no *Shigella* cases were detected by GPDS in East Asia during this time period. Data behind this figure are displayed in S1 Table.

*Shigella* AFs tended to increase with increasing age groups, indicating that *Shigella* was responsible for a greater percentage of diarrhea cases in older children than in younger children (Fig 2; S2 Table). Overall, 19.5% (95% CI: 15.7, 23.7) of hospitalized diarrhea among children aged 36-59 months was attributable to *Shigella*, and this was the age group with the highest *Shigella* AF in each geographic region except for Eastern Europe, where all AFs were low but the highest *Shigella* AF was among children aged 24-35 months (2.3%; 95% CI: 1.6%, 3.1%). The highest *Shigella* AF among the youngest children aged <6 months was in Central America (13.7%; 95% CI: 8.3%, 20.1%).

**Fig 2.**
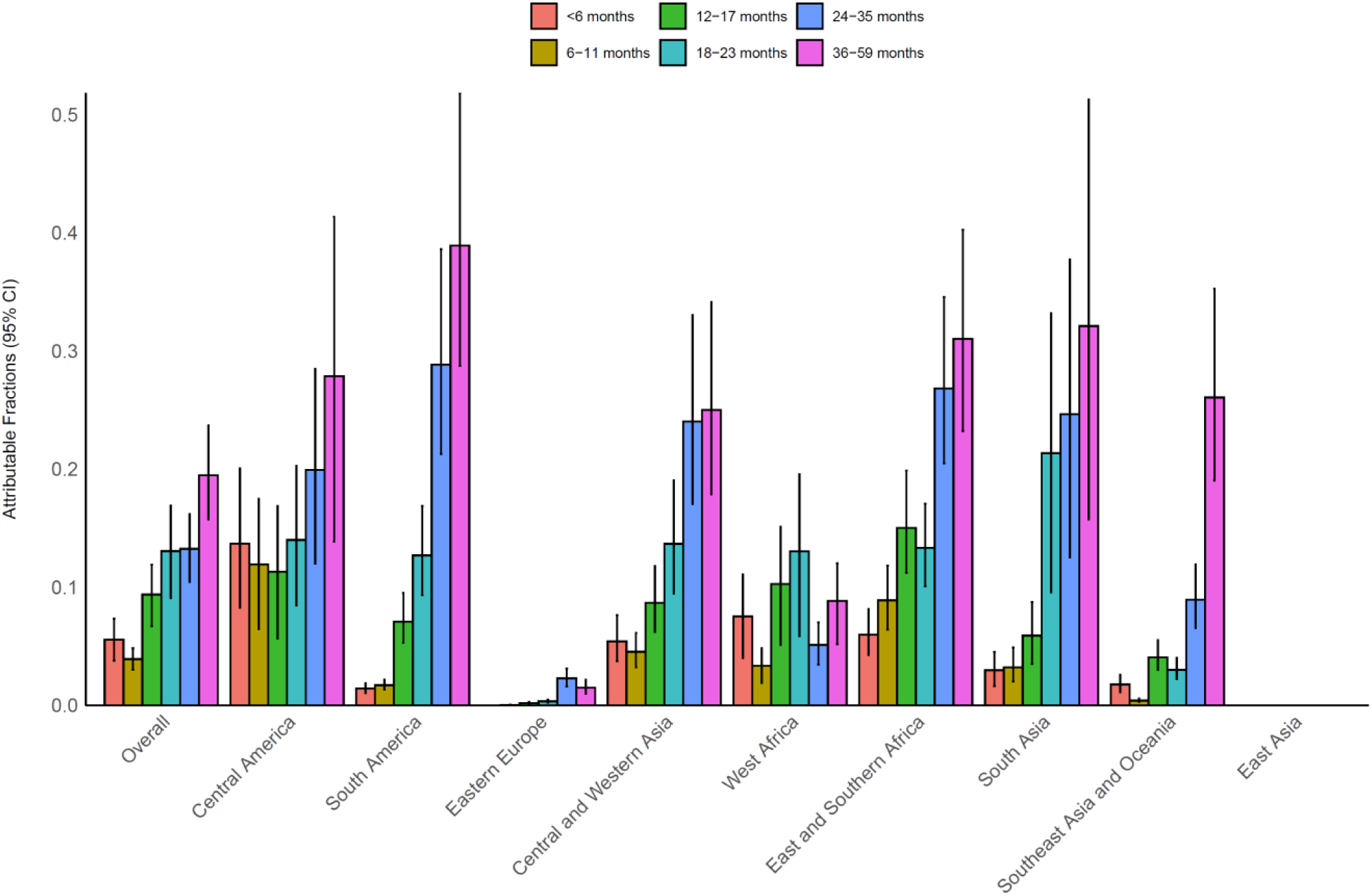
Attributable fractions of hospitalized diarrhea due to *Shigella* in children aged <5 years in 2017-2022 in the Global Pediatric Diarrhea Surveillance network by geographic region and age group. Within each grouping and pathogen, the attributable fractions were weighted by the site-level attributable incidence of hospitalized diarrhea. Attributable fractions are expressed as a percent. No bars are included for East Asia, as no *Shigella* cases were detected by GPDS in East Asia during this time period. Data behind this figure are displayed in S2 Table.

Though the overall *Shigella* AFs remained fairly stable from 2017 to 2022, the estimated attributable incidence of diarrhea hospitalizations due to *Shigella* decreased from 1.27 (95% CI: 0.95, 1.64) per 1,000 child years in 2017-2018 to 0.88 (0.67, 1.10) in 2021-2022 (Table 3). *Shigella* incidence was highest in Western Africa (decreasing from 4.18 in 2017-2018 to 1.38 in 2021-2022) and East and Southern Africa (decreasing from 3.85 in 2017-2018 to 3.33 in 2021-2022). A decrease in incidence was observed during this time period in all regions except for in Eastern Europe, where incidence remained stable and low (≤0.02), in Central and Western Asia, where incidence peaked in 2019-2020 at 0.29 (0.19, 0.42), and in South Asia, where incidence increased slightly from 0.62 (0.26, 1.02) in 2017-2018 to 0.73 (0.33, 1.10) in 2021-2022. Whether the changes in estimated *Shigella* incidence over time were driven by changes in all-cause diarrhea incidence, changes in *Shigella* AF, or both, varied by region (S1 Fig).

**Table 3.** Attributable incidence of diarrhea hospitalizations due to *Shigella* per 1,000 child years by geographic region in the Global Pediatric Diarrhea Surveillance network, 2019-2022.

| Geographic Region | Attributable Incidence (95% Confidence Intervals) |  |  |
| --- | --- | --- | --- |
|  | 2017-2018 | 2019-2020 | 2021-2022 |
| Overall | 1.27 (0.95, 1.64) | 0.87 (0.67, 1.07) | 0.88 (0.67, 1.10) |
| Central America | 0.32 (0.18, 0.47) | 0.21 (0.13, 0.29) | 0.17 (0.11, 0.23) |
| South America | 0.28 (0.20, 0.37) | 0.26 (0.19, 0.36) | 0.18 (0.13, 0.24) |
| Eastern Europe | 0.02 (0.01, 0.02) | 0.01 (0.01, 0.02) | 0.02 (0.02, 0.03) |
| Central and Western Asia | 0.12 (0.08, 0.16) | 0.29 (0.19, 0.42) | 0.19 (0.13, 0.28) |
| West Africa | 4.18 (2.36, 6.41) | 2.01 (1.04, 2.93) | 1.38 (0.72, 2.01) |
| East and Southern Africa | 3.85 (2.84, 5.14) | 3.08 (2.25, 4.06) | 3.33 (2.36, 4.49) |
| South Asia | 0.62 (0.26, 1.02) | 0.60 (0.35, 0.89) | 0.73 (0.33, 1.10) |
| Southeast Asia and Oceania | 0.63 (0.45, 0.89) | 0.33 (0.23, 0.51) | 0.57 (0.34, 0.89) |
| East Asia | 0.00 (0.00, 0.00) | 0.00 (0.00, 0.00) | 0.00 (0.00, 0.00) |

*S. sonnei* was identified in 34.8% of typeable *Shigella* cases globally (Fig 3). *S. flexneri* 2a and *S. flexneri* 1b were the next most common serotypes, representing 24.2% and 9.9% of typeable *Shigella* cases, respectively. All other *S. flexneri* serotypes were present to some degree, each representing less than 8% of overall cases. Serotype distributions in West Africa, East and Southern Africa, and South Asia were relatively consistent with the overall distribution, although *S. flexneri* 2a and 2b were more prevalent in West Africa and *S. flexneri* 2a had a lower prevalence in East and Southern Africa than overall. In Central America and Southeast Asia and Oceania, *S. sonnei* was identified in most cases, with smaller contributions from other serotypes. In two regions, *S. flexneri* 2a was more prevalent than *S. sonnei*, comprising 69.9% of cases in Central and Western Asia and 43.2% of cases in South America.

**Fig 3.**
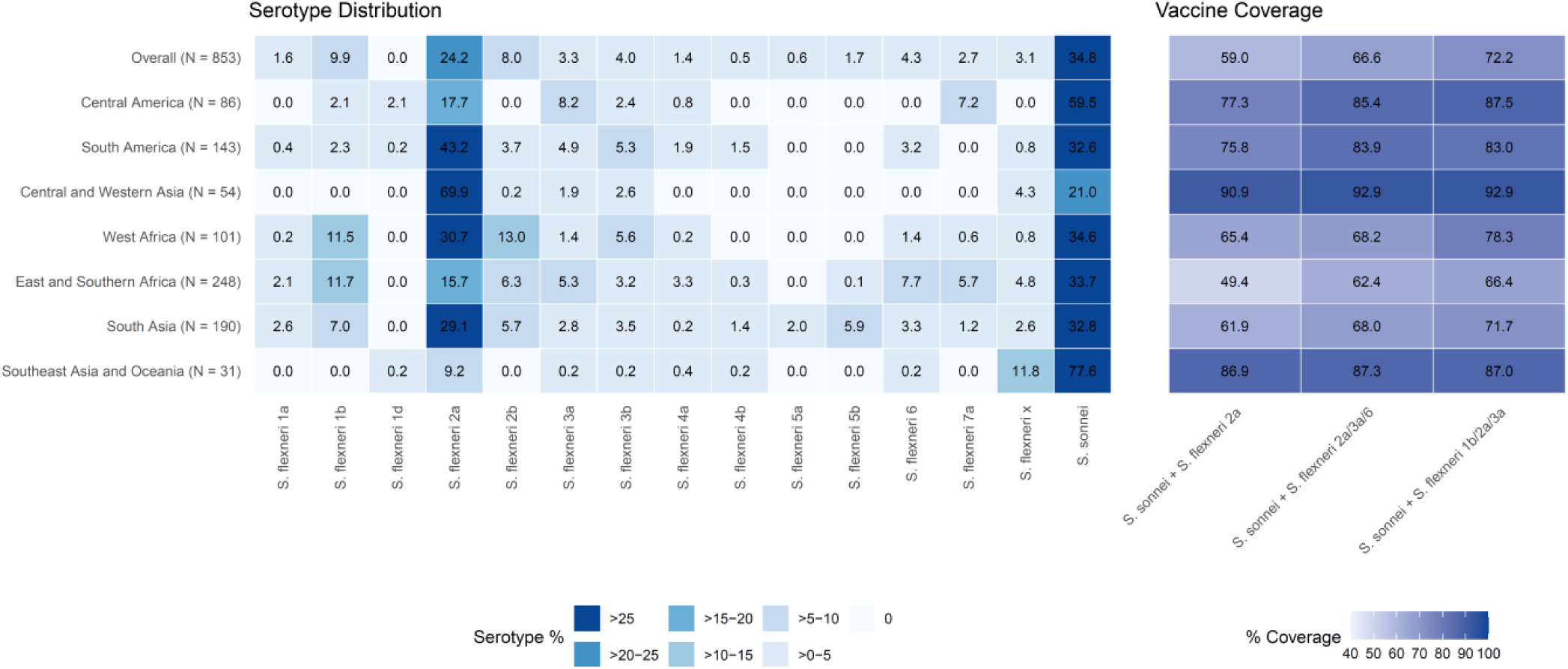
*Shigella sonnei and Shigella flexneri* serotype distributions and estimated homotypic vaccine coverage by geographic region in the Global Pediatric Diarrhea Surveillance network, 2019-2022. The number of typeable *Shigella* cases from each geographic region is indicated in parentheses. Estimates are weighted by patient-level sampling fractions and the estimated annual number of *Shigella* hospitalizations for each country, with surveillance episodes pooled across sites within multi-site countries. Only regions with ≥10 successfully typed specimens are shown.

**Fig 4.**
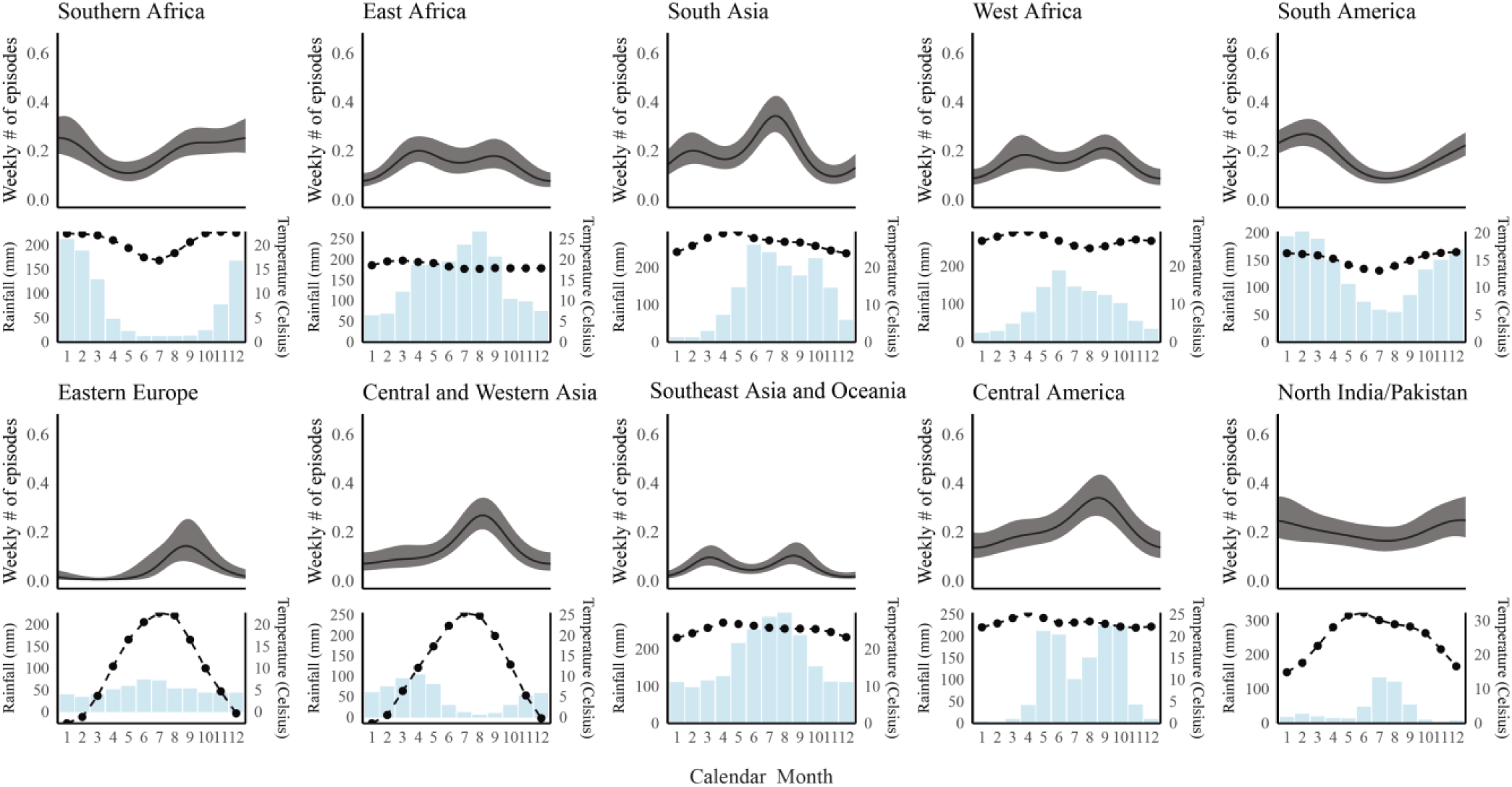
Seasonality of *Shigella-*attributable cases of hospitalized diarrhea among children aged <5 years in 2017-2022 in the Global Pediatric Diarrhea Surveillance network both overall and by geographic grouping. To evaluate for seasonal trends in pathogen-specific diarrhea, we used modified geographic groupings to cluster sites with similar latitudes within each geographic region.

Globally, a bivalent vaccine targeting *S. sonnei* and *S. flexneri* 2a would protect against an estimated 59.0% of typeable *Shigella* cases, while a quadrivalent vaccine additionally targeting *S. flexneri* 3a and 6 would cover an estimated 66.6% of cases and a quadrivalent vaccine covering *S. sonnei* + *S. flexneri* 1b, 2a, 3a would cover an estimated 72.2% of cases (Fig 3). Estimated vaccine coverage of circulating serotypes differed by region (Fig 3) and by surveillance site (S2 Fig). All three of these vaccines would have targeted almost all cases in regions such as Central and Western Asia (range: 90.9-92.9%) or Southeast Asia and Oceania (range: 86.9-87.3%). East and Southern Africa had the lowest estimated vaccine coverage: only 49.4% for the bivalent vaccine and 62.4-66.4% for the quadrivalent vaccines. Both quadrivalent vaccines had similar estimated vaccine coverage in each geographic region, with the largest difference between the vaccines occurring in West Africa, where an estimated 68.2% of typeable *Shigella* cases would have been covered by *S. sonnei* + *S. flexneri* 2a, 3a, 6 and 78.3% by *S. sonnei* + *S. flexneri* 1b, 2a, 3a.

In terms of clinical characteristics, *Shigella*-attributable cases were more likely to present with bloody diarrhea as compared to diarrhea cases where *Shigella* was not the etiology (risk ratio [RR]:3.44; 95% CI: 2.78, 4.26; Table 4). Additionally, *Shigella*-attributable cases had increased maximum daily episodes of diarrhea and decreased maximum daily episodes of vomiting and diarrhea severity score as compared to children without *Shigella.* Within *Shigella* cases, those typed as *S. flexneri* were the most likely to present with bloody diarrhea (RR 5.13; 3.94, 6.67). Otherwise, clinical characteristics were similar among *Shigella* species.

**Table 4.** Clinical characteristics of children with *Shigella*-attributable diarrhea in the Global Pediatric Diarrhea Surveillance network, 2019-2022.

| Clinical characteristics<sup>a</sup> | All *Shigella*-attributable diarrhea | *S. flexneri* | *S. sonnei* | Untyped *Shigella*<sup>b</sup> |
| --- | --- | --- | --- | --- |
| **Relative risks** |  |  |  |  |
| Vomiting (97.2) | 0.92 (0.84, 1.01) | 0.88 (0.77, 1.02) | 1.03 (0.87, 1.22) | 0.89 (0.76, 1.04) |
| Acute diarrhea (89.7) | 0.99 (0.91, 1.07) | 0.99 (0.88, 1.12) | 0.97 (0.83, 1.13) | 0.98 (0.86, 1.13) |
| Bloody diarrhea (93.7) | 3.44 (2.78, 4.26) | 5.13 (3.94, 6.67) | 1.93 (1.23, 3.03) | 2.78 (1.91, 4.04) |
| Any dehydration (65.1) | 0.92 (0.83, 1.03) | 0.90 (0.76, 1.06) | 0.97 (0.80, 1.16) | 0.95 (0.79, 1.13) |
| Severe dehydration (65.1) | 0.94 (0.84, 1.06) | 0.94 (0.79, 1.11) | 1.03 (0.85, 1.25) | 0.88 (0.72, 1.07) |
| Received IV rehydration (74.7) | 0.98 (0.89, 1.08) | 0.98 (0.85, 1.14) | 1.01 (0.84, 1.21) | 0.95 (0.8, 1.12) |
| **Mean differences** |  |  |  |  |
| Maximum daily episodes of diarrhea (98.2) | 0.61 (0.33, 0.89) | 1.19 (0.78, 1.59) | 0.53 (0, 1.05) | -0.11 (-0.57, 0.35) |
| Maximum daily episodes of vomiting (96.3) | -0.59 (-0.84, -0.34) | -0.76 (-1.13, -0.40) | -0.33 (-0.80, 0.15) | -0.47 (-0.88, -0.05) |
| Diarrhea severity score<sup>c</sup> (54.1) | -0.43 (-0.70, -0.15) | -0.32 (-0.73, 0.09) | -0.11 (-0.59, 0.38) | -0.58 (-1.03, -0.13) |

For all estimates, the clinical characteristics were modeled as a function of the *Shigella* attributable fraction for each episode, such that the estimate represents the risk or mean difference for diarrhea maximally attributable to *Shigella* compared to episodes that were not attributable to *Shigella*, and the 95% confidence interval is presented in parentheses. Differences in continuous metrics were assessed using linear regression. For binary variables, Poisson regression with a log link was used to estimate relative risks. For the type-specific estimates (final three columns), episodes assigned to the alternative types were excluded from the model.
<sup>a</sup>The number in parentheses after each clinical characteristic represents the proportion of episodes with available data.
<sup>b</sup>Untyped *Shigella* could include *S. boydii*, *S. dysenteriae*, and enteroinvasive *E. coli* as well as potentially unidentified *S. sonnei* and *S. flexneri* serotypes.
<sup>c</sup>The diarrhea severity score had a maximum possible score of 16 (19).

## Discussion

Through sustained surveillance of children with diarrhea requiring hospitalization in a wide range of LMICs over a six-year period, this analysis demonstrates the ongoing importance of *Shigella* as an etiology of severe pediatric diarrhea, comprising around 10% of diarrheal hospitalizations. This analysis also substantially increases the available data on the distribution of *Shigella* species and serotypes to aid in vaccine development and provides additional data on the age distribution, seasonality, and clinical presentation of severe shigellosis. Importantly, these data are specifically from children aged <5 years in LMICs, a population which WHO has identified as the priority target for a *Shigella* vaccine (14,20).

The finding that *Shigella* was a major cause of hospitalized diarrhea in children in LMICs, especially after rotavirus vaccine introduction, is consistent with other studies that have generally studied a broader spectrum of diarrhea severity (4,25). Geographically, *Shigella* was overrepresented in diarrheal cases in regions such as East and Southern Africa, South America, and South Asia, making these regions important areas to target with vaccination and treatment efforts. East and Southern Africa and South Asia may be especially important regions for intervention as they have been seeing steady increases in *Shigella* attributable fractions over the past several years. While *Shigella* caused a higher proportion of hospitalizations in older children, nearly a third (and more than 40% in Central America) of cases occurred by 12 months of age. This suggests that vaccines should target younger infants if technically feasible (11,26).

Through direct identification of *S. sonnei* and *flexneri* serotypes from stool without the relying on isolates, this analysis provides data on the distribution of these *Shigella* species and serotypes that are prioritized for vaccine development without any potential bias due to species-specific culture sensitivity (15,16). These data suggest considerable regional variation in homotypic coverage of vaccines in current clinical development (11). For example, a bivalent vaccine against *S. sonnei* and *S. flexneri 2a* would directly target almost all cases in some areas but less than half of cases in others, which in the absence of significant heterotypic protection may be better served by introduction of a quadrivalent vaccine. These broadly representative data can aid in vaccine prioritization, both for clinical development and for eventual vaccine deployment.

The association of *Shigella* cases in this cohort with bloody diarrhea and not with vomiting is consistent with the described clinical presentation of pediatric shigellosis (8). *Shigella* was slightly less severe than other causes of hospitalized diarrhea, a finding that might have been driven by the older age distribution. Comparison of clinical characteristics within *Shigella* species was also largely as expected, with *S. flexneri* being more strongly associated with bloody diarrhea. Otherwise, species appear to be similar in terms of presentation and severity, supporting existing characterizations. Recognizing that *S. sonnei* is less likely to present with blood is important, because current WHO treatment guidelines recommend antibiotics only for children with bloody diarrhea (27). Strategies to target acute watery shigellosis for antibiotic therapy require further evaluation (28).

*Shigella* is classically thought to have strong seasonal variation, with a higher incidence during warm, rainy seasons (4,29). This significant seasonality largely proved true in the GPDS cohort, although with regional differences in the level of variation and in the association with rainfall. Knowing the seasonal variation may may be useful in guiding empiric treatment of diarrheal cases, especially in seasons with high *Shigella* prevalence. A comparison of seasonality between *Shigella* species would also be both interesting and useful in guiding treatment decisions, but the size of this cohort did not allow for seasonality modeling at that level.

GPDS is a strong platform to investigate *Shigella* epidemiology among children aged <5 years in LMICs, covering a wide range of geographic settings, enrolling and testing large numbers of children, and using comprehensive culture-independent diagnostic tools to identify *Shigella* and its serotypes (6,19). However, the cohort size is insufficient for statistical modeling of species- or serotype-specific seasonality and potentially for distinguishing differences in clinical characteristics. Particularly as some clinical variables were not complete for all enrolled children, especially dehydration and whether rehydration therapy was given. Additionally, GPDS has limited representation in the Middle East, North Africa, and East Asia, potentially challenging the representativeness of this analysis for those geographies. Though the GPDS TAC cards included ten targets to identify *Shigella* species and serotypes, not identify all *Shigella* species and serotypes could be identified, particularly in some regions with a higher proportion of untyped *Shigella.* Additionally, because catchment area and healthcare utilization estimates are not available for the GPDS sites, we rely on extrinsic estimates of country-level diarrhea incidence to aggregate AF estimates between sites as well as for all incidence estimates (19,22). Finally, these analyses of potential vaccine coverage for three *Shigella* vaccines in development exclusively estimate homotypic vaccine coverage; any heterotypic vaccine coverage would increase these estimates.

These findings support the role of *Shigella* as an important cause of the most severe subset of diarrhea in young children. Understanding when, where, and how *Shigella* is causing illness is vital for appropriately developing interventions. This analysis of GPDS data details substantial geographic variation in both *Shigella* epidemiology and estimated homotypic *Shigella* vaccine coverage, indicating that a region-specific approach to vaccination and treatment guidelines is likely needed. More effectively targeted *Shigella* prevention and treatment strategies would have far-reaching health benefits for pediatric populations in LMICs.

## Data Availability

Aggregate GPDS data is available online at: https://immunizationdata.who.int/global?topic=Rotavirus-and-pediatric-diarrhea-surveillance-data. De-identified participant-level data used in these analyses will be made available by WHO upon request to qualified researchers, after approval of a proposal submitted to and signing of a WHO data sharing agreement.

## Acknowledgements

We would like to thank the sentinel surveillance hospitals, national laboratories, and staff and the country Ministries of Health for supporting and maintaining surveillance at the country level, as well as the WHO Country Offices. The findings and conclusions of this report are those of the authors and do not necessarily represent the official position of the World Health Organization.

## Supplemental Material

**S1 Table.** Attributable fractions of hospitalized diarrhea due to *Shigella* in children aged <5 years in 2017-2022 in the Global Pediatric Diarrhea Surveillance network both overall and by geographic region (Underlying data for Figure 1).

| Geographic Region | Attributable Fractions (95% Confidence Intervals) |  |  |
| --- | --- | --- | --- |
|  | 2017-2018 | 2019-2020 | 2021-2022 |
| Overall | 10.6 (8.0, 13.6) | 8.7 (6.7, 10.7) | 10.1 (7.7, 12.6) |
| Central America | 17.3 (9.8, 25.9) | 12.0 (7.9, 17.1) | 9.5 (6.4, 13.2) |
| South America | 12.2 (8.9, 16.4) | 13.9 (9.9, 19.0) | 12.1 (9.0, 16.2) |
| Eastern Europe | 1.2 (0.9, 1.6) | 0.8 (0.5, 1.2) | 1.1 (0.8, 1.7) |
| Central and Western Asia | 10.5 (7.4, 14.3) | 13.3 (8.7, 19.5) | 9.6 (6.3, 14.0) |
| West Africa | 9.6 (5.4, 14.7) | 5.6 (2.9, 8.2) | 4.7 (2.5, 6.9) |
| East and Southern Africa | 14.3 (10.5, 19.0) | 15.4 (11.0, 20.4) | 18.4 (13.1, 24.8) |
| South Asia | 12.4 (5.2, 20.5) | 12.5 (7.2, 18.5) | 16.8 (7.8, 25.5) |
| Southeast Asia and Oceania | 6.1 (4.3, 8.6) | 2.9 (2.0, 4.5) | 5.4 (3.2, 8.5) |
| East Asia | 0.0 (0.0, 0.0) | 0.0 (0.0, 0.0) | 0.0 (0.0, 0.0) |

**S2 Table.** Attributable fractions of hospitalized diarrhea due to *Shigella* in children aged <5 years in 2017-2022 in the Global Pediatric Diarrhea Surveillance network by geographic region and age group (Underlying data for Figure 2).

| Geographic Region | Attributable Fractions (95% Confidence Intervals) |  |  |  |  |  |
| --- | --- | --- | --- | --- | --- | --- |
|  | <6 months | 6-11 months | 12-17 months | 18-23 months | 24-35 months | 36-59 months |
| Overall | 5.6 (3.8, 7.3) | 3.9 (3.0, 4.8) | 9.4 (6.7, 11.9) | 13.0 (9.1, 16.9) | 13.2 (10.5, 16.2) | 19.5 (15.7, 23.7) |
| Central America | 13.7 (8.3, 20.1) | 11.9 (6.5, 17.5) | 11.3 (5.7, 16.9) | 14.0 (8.4, 20.3) | 19.9 (12.0, 28.5) | 27.9 (13.9, 41.4) |
| South America | 1.4 (1.0, 1.9) | 1.7 (1.3, 2.2) | 7.1 (5.3, 9.5) | 12.7 (9.3, 16.9) | 28.8 (21.3, 38.6) | 38.9 (28.7, 51.9) |
| Europe | 0.0 (0.0, 0.0) | 0.0 (0.0, 0.1) | 0.2 (0.1, 0.3) | 0.3 (0.2, 0.5) | 2.3 (1.6, 3.1) | 1.5 (1.0, 2.2) |
| Central Asia | 5.4 (3.7, 7.6) | 4.5 (3.2, 6.1) | 8.7 (6.2, 11.8) | 13.7 (9.5, 19.0) | 24.0 (17.0, 33.0) | 25.0 (17.9, 34.1) |
| West Africa | 7.5 (4.0, 11.1) | 3.3 (1.9, 4.8) | 10.3 (5.1, 15.1) | 13.0 (5.8, 19.6) | 5.1 (3.4, 7.0) | 8.8 (5.2, 12.0) |
| East Africa | 6.0 (4.3, 8.2) | 8.9 (6.4, 11.8) | 15.0 (11.2, 19.9) | 13.3 (10.1, 17.1) | 26.8 (20.5, 34.6) | 31.0 (23.2, 40.3) |
| South Asia | 3.0 (1.6, 4.5) | 3.2 (2.0, 4.9) | 5.9 (3.5, 8.8) | 21.4 (9.6, 33.2) | 24.6 (12.5, 37.7) | 32.1 (15.7, 51.3) |
| Southeast Asia and Oceania | 1.8 (1.1, 2.6) | 0.4 (0.3, 0.6) | 4.1 (3.0, 5.5) | 3.0 (2.2, 4.0) | 8.9 (6.5, 11.9) | 26.1 (19.0, 35.3) |
| East Asia | 0.0 (0.0, 0.0) | 0.0 (0.0, 0.0) | 0.0 (0.0, 0.0) | 0.0 (0.0, 0.0) | 0.0 (0.0, 0.0) | 0.0 (0.0, 0.0) |

**S1 Fig.**
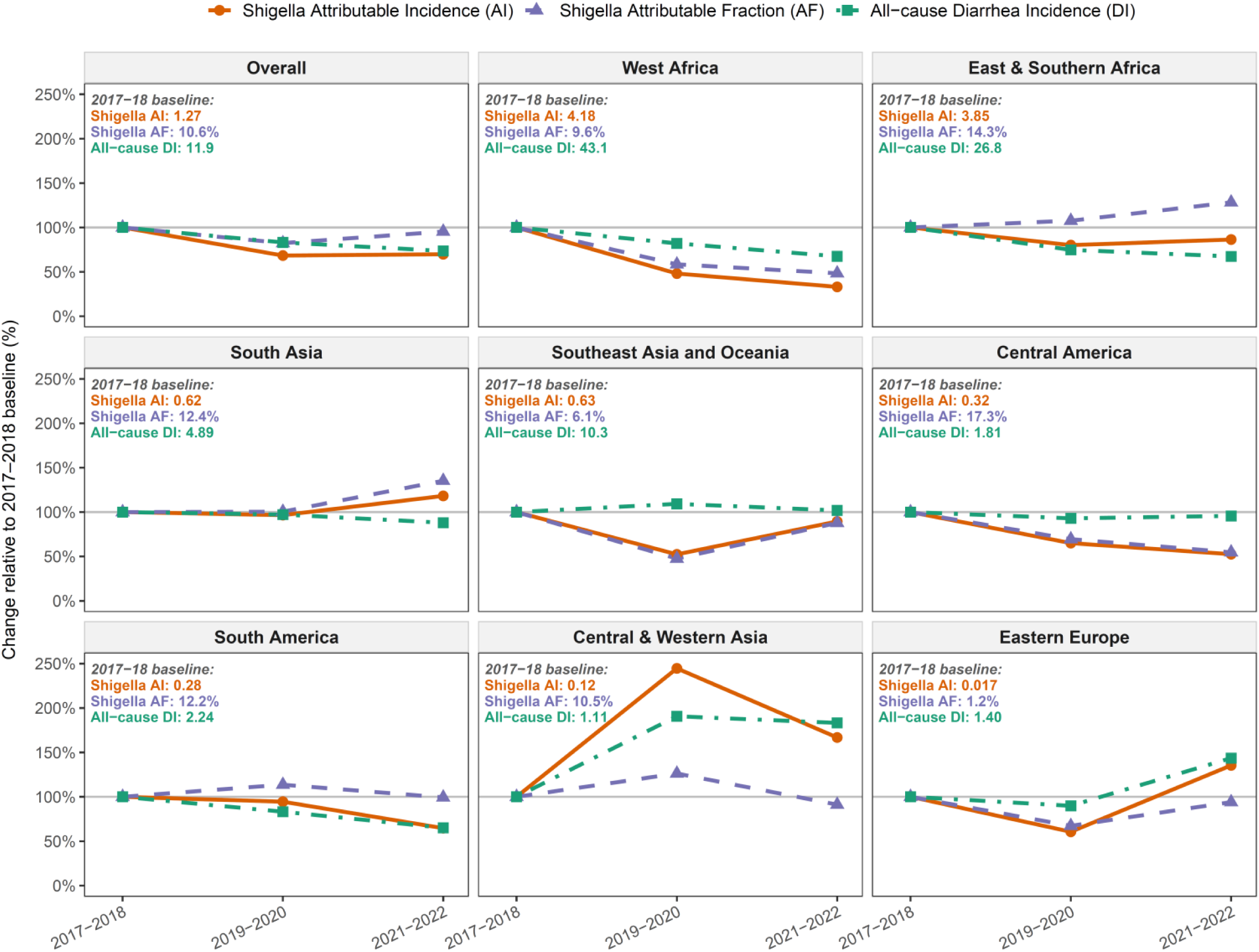
Change in *Shigella* attributable fraction, attributable incidence, and all-cause diarrhea incidence from 2017-2018 (baseline) to 2019-2020 and 2021-2022.

**S2 Fig.**
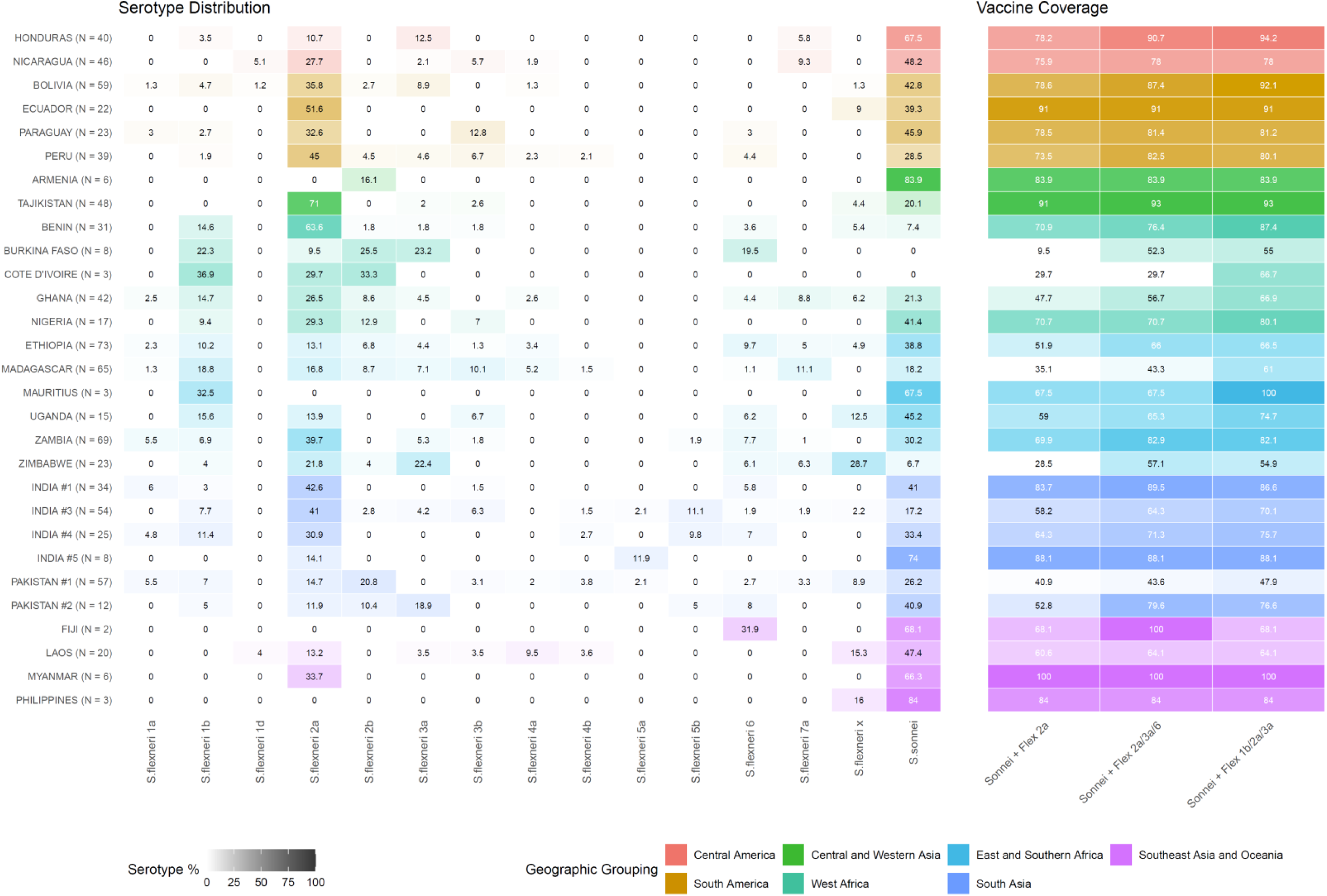
*Shigella sonnei* and *Shigella flexneri* serotype distributions and estimated homotypic vaccine coverage by surveillance site in the Global Pediatric Diarrhea Surveillance network, 2019-2022. The number of typeable *Shigella* cases from each site is indicated in parentheses. Estimates are weighted by patient-level sampling weights.

